# Ready for Impact? A validity and feasibility study of instrumented mouthguards (iMGs)

**DOI:** 10.1101/2022.01.28.22270039

**Authors:** Ben Jones, James Tooby, Dan Weaving, Kevin Till, Cameron Owen, Mark Begonia, Keith Stokes, Steve Rowson, Gemma Phillips, Sharief Hendricks, Éanna Falvey, Marwan Al-Dawoud, Gregory Tierney

## Abstract

**Objectives:** Determine the validity and feasibility of current Instrumented mouthguards (iMGs) and associated systems.

**Methods:** Phase 1; Four iMG systems (Football Research Inc [FRI], HitIQ, ORB, Prevent) were compared against dummy headform laboratory criterion standards (25, 50, 75, 100 *g*). Phase 2; Four iMG systems were evaluated for on-field validity of iMG-triggered events against video-verification to determine true-positives, false-positives and false-negatives (20 ± 9 player matches per iMG). Phase 3; Four iMG systems were evaluated by eighteen rugby players, for perceptions of *fit, comfort* and *function*. Phase 4; Three iMG systems (FRI, HitIQ, Prevent) were evaluated for practical feasibility (system usability scale; SUS) by four practitioners.

**Results:** Phase 1; Total concordance correlation coefficient was 98.3%, 95.3%, 42.5% and 97.9% for FRI, HitIQ, ORB and Prevent. Phase 2; Different on-field kinematics were observed between iMGs. Positive predictive values were 0.98, 0.90, 0.53 and 0.94 for FRI, HitIQ, ORB and Prevent. Sensitivity values were 0.51, 0.40, 0.71 and 0.75 for FRI, HitIQ, ORB and Prevent. Phase 3; player perceptions of *fit, comfort* and *function* were 77%, 6/10, 55% for FRI, 88%, 8/10, 61% for HitIQ, 65%, 5/10, 43% for ORB, and 85%, 8/10, 67% for Prevent. Phase 4; SUS was 51.3-50.6/100, 71.3-78.8/100, and 83.8-80.0/100 for FRI, HitIQ, and Prevent.

**Conclusion:** This study shows that differences between current iMG systems exist. Sporting organisations can use these findings to ensure accurate head acceleration event data are obtained and system adoption is optimized, to support player welfare initiatives directly related to long-term brain health.

## INTRODUCTION

Instrumented mouthguards (iMGs) have potential to quantify head acceleration events (HAEs) in sport. HAEs can occur due to direct impacts to the head or inertial head loading from impacts to the body. iMGs can provide data to quantify the cumulative HAE experienced by players and also establish the biomechanical mechanisms of injury for concussion, which are a concern for sports.[1] Limited validation and feasibility studies exist that evaluate iMGs and their associated systems,[2,3] thus their appropriateness for both research and practice is unknown. Prior to the application and adoption of iMGs in sport, their validity and feasibility require investigation to enable sporting organisations, clinicians, and the scientific community to be aware of the strengths and limitations of iMGs. Valid iMG data can inform player welfare, safety, and medical initiatives, directly related to long-term brain health.[4].

This four-phase study[4] aimed to determine; Phase 1) the validity of iMG kinematic magnitude measures against laboratory criterion standards, Phase 2) the on-field validity of iMGs via video-verification, Phase 3) iMG feasibility evaluated via player perceptions of *fit, function* and *comfort*, and Phase 4) practical feasibility of iMG systems from a practitioner perspective.

## METHODS

### Study Design

The four-phase study evaluated four currently available iMG systems, consistent with the published protocol.[4] Phase 1 validated the kinematic measures of iMGs against laboratory criterion standards.[2,3] Phase 2 evaluated the on-field validity of iMG-triggered events by identifying true-positives, false-positives and false-negatives via video-verification from 80 rugby league player matches (20 ± 9 player matches per iMG company). Phase 3 evaluated the *fit, comfort* and *function*,[5] of iMGs from a player’s perspective, and Phase 4 evaluated the practical feasibility of the iMG system from a practitioner’s perspective[6,7] using questionnaires.

### Instrumented Mouthguard Recruitment

iMG companies were invited to provide 43 iMGs (*n* = 3 Phase 1, *n* = 20 Phase 2, *n* = 20 Phase 3) and associated systems free of charge. The protocol[4] was shared, and companies were informed that the findings would help determine the appropriateness of iMGs for the TaCKLE (Tackle and Contact Kinematic, Load and Exposure) project.[8] Six iMG companies were initially approached, one declined, and one withdrew prior to data collection. Four companies provided iMGs and associated systems (Table 1).

**Table 1.**
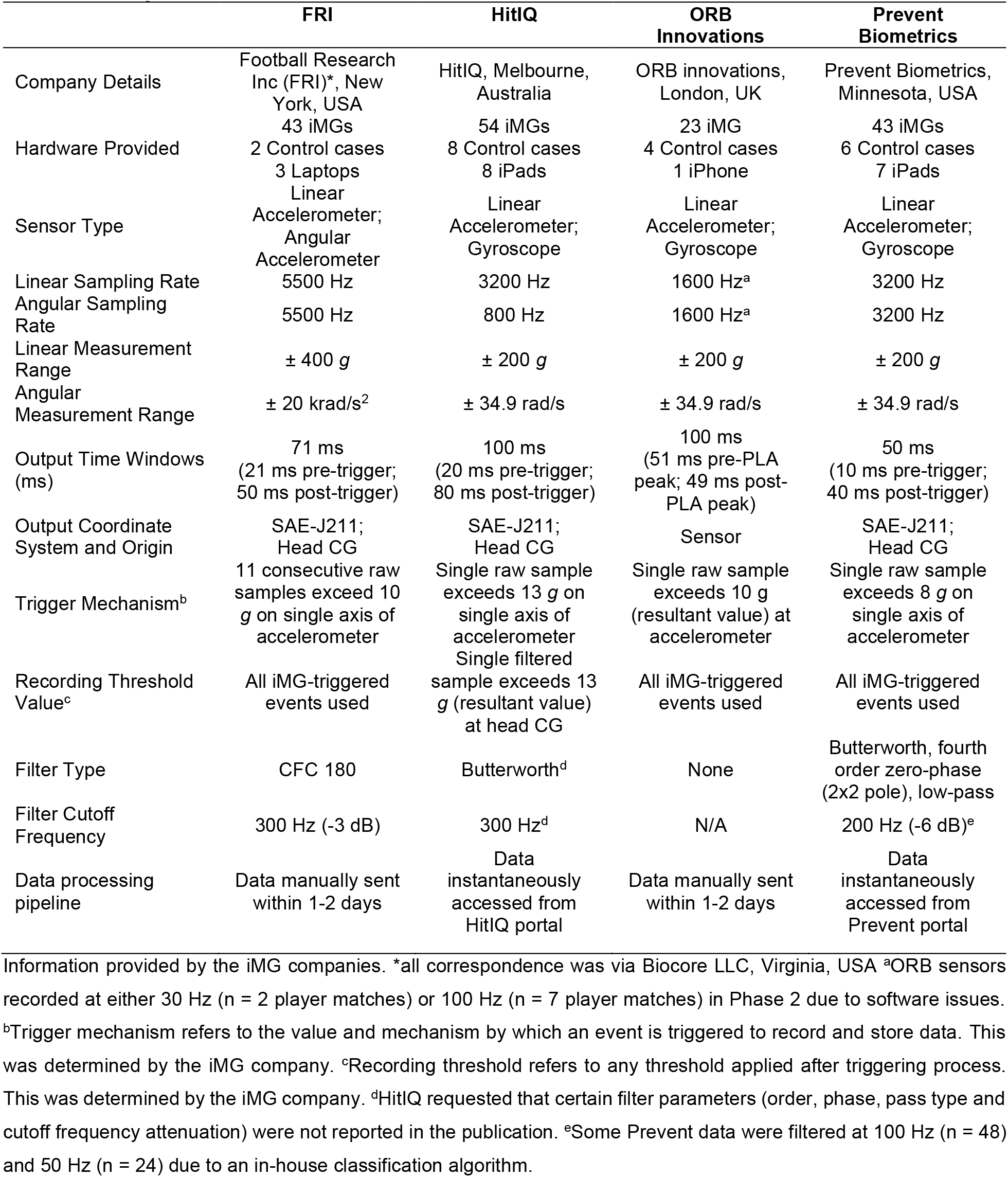
Hardware provided for the study, and product specification of iMGs and associated systems.

### Phase 1 – Laboratory Validation of Kinematic Measures

In-laboratory testing to evaluate the validity of iMG kinematic magnitudes was conducted at the Virginia Tech Helmet Lab (USA),[2] using a bareheaded dummy headform (Figure 1a)[9,10] impacted at various locations, impact magnitudes (target linear head accelerations 25, 50, 75, and 100 *g*) and durations (rigid; nylon, 25 mm thickness, and padded; vinyl-nitrile foam, 40 mm thickness) using a pendulum impactor (127 mm diameter) (Figure 1b) as previously described.[4] Two tests were conducted at each configuration and three iMGs were tested for each iMG company (Figure 1c). Reference kinematics were measured at the headform centre of gravity with an instrumentation package consisting of three linear accelerometers (Endevco 7264b-2000; Meggitt Orange County, Irvine, CA) and a tri-axial angular rate sensor (DTS ARS3 Pro 18k; Diversified Technical Systems, Seal Beach, CA), recording at 20 kHz, and filtered using a CFC 1000 and CFC 155. Custom-fit iMGs were mounted inside the headform as previously described.[4] HitIQ repeated testing, due to an issue with one test configuration producing high errors during the first round. HitIQ believed this was due to the use of their non-research data portal and/or experimental error. HitIQ implemented a firmware update and provided access to a research portal for the second round of testing. Football Research Inc [FRI] conducted three additional tests at the lowest speed test configuration. During the first round of testing, this configuration did not record as it was below the specific set iMG-trigger threshold. ORB updated the impact data analysis to account for an issue involving the impact detection algorithm and the iMGs two onboard accelerometers. ORB noted their algorithm sometimes saved data from the iMGs low-instead of high range accelerometer (±16 *vs*. ±200 *g*). Prevent had previously been tested in this laboratory using the same methodology.[2] HitIQ undertook pilot testing, independent of the research team in this laboratory prior to the study. Peak resultant linear acceleration (PLA), peak resultant angular velocity (PAV), and peak resultant angular acceleration (PAA) were recorded.[4]

**Figure 1.**
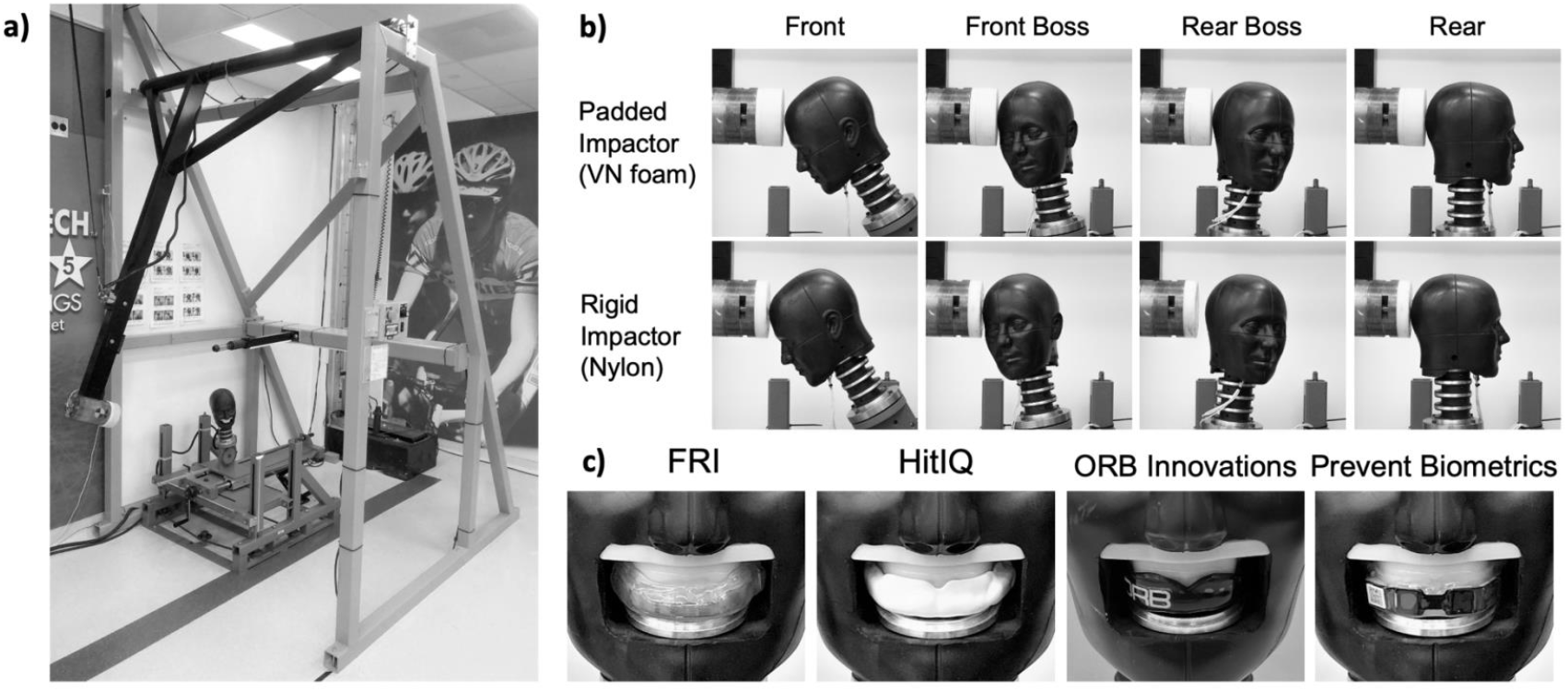
**(a) Experimental set-up of pendulum impactor to simulate bareheaded impacts to the dummy headform. (b) Padded (vinyl-nitrile) and rigid (nylon) impactor to the bareheaded dummy headform at the front, front boss, rear boss, and rear locations of the headform. (c) The custom-fit iMG mounted inside the headform with detachable 3D-printed detention**.

### Phase 2 – On-Field Validity

Fifty-one male rugby league players from five Super League academies underwent 3D dental scans, performed by an experienced dentist, and reviewed by a third-party dentist with experience in iMG manufacture. Each company manufactured the custom-fit iMGs, for two teams, based on dental details received via a standard language tessellation file. HitIQ provided 11 additional iMGs at the request of the research team to increase match observations. Due to injuries, suspensions, and non-compliance of iMG wearing eighty player matches were included (*n* = players wearing iMG/players received dental scans [player matches]; FRI *n* = 10/20 [17], HitIQ *n* = 18/31 [28], ORB *n* = 7/20 [9], and Prevent *n* = 12/20 [27]). For ORB, manufacturing delays reduced data collection, and battery issues resulted in 6/9 player matches having partial data collected (mean duration = 30:22 ± 09:51 mins). One player removed their Prevent iMG five minutes into a match, so data were only partially captured for this match.

All data collection and management were undertaken by the research team as per manufacturer instructions. Events recorded by iMGs were time-synchronized with high-quality video footage of match-play recorded by the home team to verify whether each iMG-triggered event (post application of companies’ recording threshold and/or HAE detection algorithm, if applicable) was associated with a HAE for the instrumented player. Companies determined the iMG recording and trigger thresholds (Table 1), as previously described.[4] iMG data were trimmed to synchronize with kick-off and 10 minutes post-match, resulting in data being recorded during the half-time period.

To determine true-positives or false-positives, a guided analysis was undertaken. iMG-triggered events were classified by a trained video analyst as true-positives, false-positives or assumed false-positives (Table 2). To determine false-negatives, an unguided analysis was undertaken. One professional rugby video analyst tracked each player wearing an iMG during matches and labelled their one-on-one shoulder tackles when a tackler.[11] The timestamp of each labelled one-on-one shoulder tackle event was cross-referenced with the iMG-triggered dataset. True-positives or false-negatives were based on whether the one-on-one shoulder tackle event timestamp matched an iMG-triggered timestamp (Table 2). Only periods of play where iMGs were collecting data were used to avoid false-negatives being labelled when iMGs were not worn or active. The collection, analysis and reporting is consistent with the published protocol.[4] Additional analysis and reporting of variables included quantification of player activities during identified false-positives, a breakdown of false-negatives, and iMG kinematic magnitudes during match-play.

**Table 2.** True-positive, false-positive, assumed false-positive, false-negative definitions during for the guided and unguided analysis (Phase 2).

|  | True-Positive | False-Positive | Assumed False-Positive<br>(Off-Camera)<br>Guided analysis | Assumed False-Positive (Inactive) | True-Positives<br>Unguided analysis | False-Negative |
| --- | --- | --- | --- | --- | --- | --- |
| iMG-triggered event | Impact reported | Impact reported | Impact reported | Impact reported | Impact reported | Impact not reported |
| Video Footage | Video footage shows a visible HAE from a contact event at an iMG-triggered event timestamp. | Video footage shows that player does not have a visible HAE from a contact event at timestamp of iMG-triggered event | iMG-triggered event occurs during active match-play whilst the instrumented player is on the field. The instrumented player is not visible on the video footage. Given the video footage is following play (i.e., the ball, tackles and carries), it is assumed this is a false-positive. | iMG-triggered event occurs during half-time or during the 10-minute period following full-time. Video footage is not available to confirm if a HAE occurred (e.g., during warm up tackles) | Instrumented player is involved in a one-on-one front on shoulder tackle, as the tackler. | Instrumented player is involved in a one-on-one front on shoulder tackle, as the tackler. |

### Phase 3 – Player feasibility (perception of *fit, comfort* and *function*)

Eighteen male rugby league players with no prior experience of iMGs, from four Super League clubs, received one iMG from each company (*n*=4), following dental scans as per Phase 2. Using a randomised cross-over design between clubs, each iMG was worn during two training sessions (>45-minutes). Within one hour following the second training session, players rated the *fit, comfort* and *function* of iMGs[5] using an online questionnaire (Qualtrics^XM^, Washington, USA). This resulted in the completion of four questionnaires for each iMG. The research team supervised players to ensure the questionnaires were completed independent of other players. *Fit, comfort* and *function* were evaluated using eight, six and four questions, answered on binary, 10- and 3-point Likert scales.[5] Results were reported consistent with the protocol.[4]

### Phase 4 – Practitioner feasibility (perception of usability)

Four physiotherapy / sports science practitioners with no prior experience of the iMG systems, from four different Super League clubs evaluated three iMG systems (HitIQ, Prevent, FRI) in a randomised cross-over design. ORB did not participate in Phase 4, at their request, due to the software requiring their interpretation and involvement. Usability was evaluated for 1) preparing iMG systems (e.g., setup, charging and deployment of the iMGs to participants) and 2) managing iMG data (e.g., extracting data to software interface, accessing information and feedback mechanisms within system). iMG companies provided an on-line ‘on-boarding’ session (observed by the research team) to familiarise practitioners with the operating procedures one day before initial use. Practitioners had access to operating procedure documentation, and support from research team/iMG companies if required. Practitioners completed the preparation and management of iMG systems on two training days. Within one hour of downloading data from second training session, practitioners completed two online System Usability Scale (SUS) questionnaires to evaluate the preparation and management of the iMG system (Qualtrics^XM^, Washington, USA) each including 10 questionnaire items combined into a single score out of 100 [6,7].

#### Data Analysis

##### Phase 1

Validity was assessed by calculating the concordance correlation coefficient (CCC) (Eq. 1), to quantify the agreement between iMGs and reference measurements.[2]

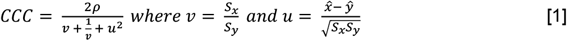

In Equation [1], ρ represents the Pearson correlation coefficient, *x* and y represent the reference and iMG measurements, respectively. *X^* and y represent the measurement means, and S_x_ and S_y_ represent the measurement standard deviations (SD). iMG output the peak resultant values for linear (e.g., PLA) and rotational (e.g., PAA) kinematic measures. After the sensor and reference measurements were recorded, both were normalized relative to the maximum reference measurement. CCC values were computed for the linear and rotational kinematic measure(s), and the combination of linear and rotational measures. The combined CCC value that accounts for peak linear and rotational acceleration represented the overall iMG in-laboratory validity. However, the ORB iMG system was unable to output PAA, and thus PAV was utilized in their combined CCC value.

##### Phase 2

Positive predictive values (PPV) (Eq. 2) were calculated for true-positives and false-positives, as well as combinations of assumed false-positives (i.e., false-positives summed).[2]

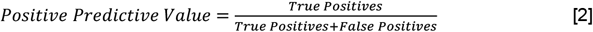

A sensitivity score was calculated (Eq 3) from the true-positive and false-negative counts for each iMG.

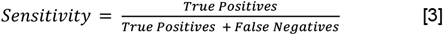

##### Phase 3

Mean (range) percentage of positive responses (i.e., ‘no’) for individual questionnaire items, evaluating *fit* and *function* were calculated. Median and interquartile ranges were calculated from the 10-point Likert questionnaire items evaluating *comfort*.

##### Phase 4

The mean ± SD of the two SUS scores (i.e., preparation and management) were calculated.

## RESULTS

Table 3 presents in-laboratory (Phase 1) and on-field (Phase 2) validity, player evaluations of *fit, comfort* and *function* (Phase 3) and practitioner SUS (Phase 4). The linear and angular kinematic values for each iMG are shown in Figure 2 (Phase 1) and Figure 3 (Phase 2). Supplementary Table 1 shows off-camera assumed false-positives and associated PPV, and a breakdown of false-positives and false-negatives. Supplementary Table 2 shows true-positives, false-positives and associated PPV for each player from Phase 2. Supplementary Table 3 shows the *fit, comfort* and *function* for each questionnaire item from Phase 3. Supplementary Figure 2 shows the linear and angular kinematic values, associated PPVs, true-positives, false-positives, and combinations of false-positives and assumed false-positives for each iMG from Phase 2.

**Table 3.** Laboratory (Phase 1) and on-field (Phase 2) validity and player (Phase 3) and practitioner (Phase 4) feasibility results.

|  | FRI | HitIQ | ORB | Prevent |
| --- | --- | --- | --- | --- |
| **Phase 1** |  |  |  |  |
| Linear CCC (%) | 97.5 | 92.1 | 32.1 | 97.2 |
| Rotational CCC (%) | 98.9 | 97.9 | 43.1 | 98.0 |
| Total CCC (%) | 98.3 | 95.3 | 42.5 | 97.9 |
| **Phase 2 (False-Positive; Guided Analysis)** |  |  |  |  |
| Player Matches (n) | 17 | 28 | 9 | 27 |
| Total Impacts (n) | 201 | 405 | 179 | 745 |
| (mean ± SD per player, per match) | (11.8 ± 10.5) | (14.5 ± 7.4) | (19.9 ± 22.8) | (27.6 ± 15.6) |
| True-Positives (n) | 162 | 245 | 66 | 666 |
| (mean ± SD per player, per match) | (9.5 ± 5.1) | (8.8 ± 5.2) | (7.3 ± 11.7) | (24.7 ± 14.6) |
| False-Positives (n) | 4 | 28 | 59 | 45 |
| (mean ± SD per player, per match) | (0.2 ± 0.4) | (1.0 ± 1.2) | (6.6 ± 7.7) | (1.7 ± 2.3) |
| True-Positives and False-Positives PPV | 0.98 | 0.90 | 0.53 | 0.94 |
| Inactive Assumed False-Positives (n) | 30 | 21 | 7 | 0 |
| (mean ± SD per player, per match) | (1.8 ± 7.0) | (0.8 ± 1.2) | (0.8 ± 2.3) | (0.0 ± 0.0) |
| True-Positives, False-Positives and Inactive Assumed False-Positives PPV | 0.83 | 0.83 | 0.50 | 0.94 |
| All Assumed False-Positives (n) | 35 | 132 | 54 | 34 |
| (mean ± SD per player, per match) | (2.1 ± 7.2) | (4.7 ± 4.9) | (6.0 ± 5.8) | (1.3 ± 2.8) |
| True-Positives, False-Positives and all Assumed False-Positives PPV | 0.81 | 0.60 | 0.37 | 0.89 |
| **Phase 2 (False-Negative; Unguided Analysis)** |  |  |  |  |
| True-Positive (n) | 47 | 49 | 20 | 82 |
| False-Negative (n) | 46 | 73 | 8 | 27 |
| Sensitivity (%) | 0.51 | 0.40 | 0.71 | 0.75 |
| **Phase 3** |  |  |  |  |
| Fit (mean % [range]) | 77 (16,100) | 88 (63,100) | 65 (22,94) | 85 (67,100) |
| Comfort (median [IQR]) | 6 (5,7) | 8 (6,8) | 5 (3,7) | 8 (7,8) |
| Function (mean % [range]) | 55 (16,100) | 61 (21,95) | 43 (11,83) | 67 (44,94) |
| **Phase 4** |  |  |  |  |
| Preparation (SUS score out of 100) | 51.3 ± 21.7 | 71.3 ± 20.7 | - | 83.8 ± 21.3 |
| Management (SUS score out of 100) | 50.6 ± 28.5 | 78.8 ± 11.1 | - | 80.0 ± 20.7 |

Note: All assumed false-positives are for summed off-camera and inactive assumed false-positives; off-camera assumed false-positives are shown in Supplementary Table 1; Preparation and Management are reported as mean and standard deviation; CCC = Concordance Correlation Coefficient; PPV = Positive Predictive Values; SUS = system usability scale; IQR = inter-quartile range.

**Figure 2.**
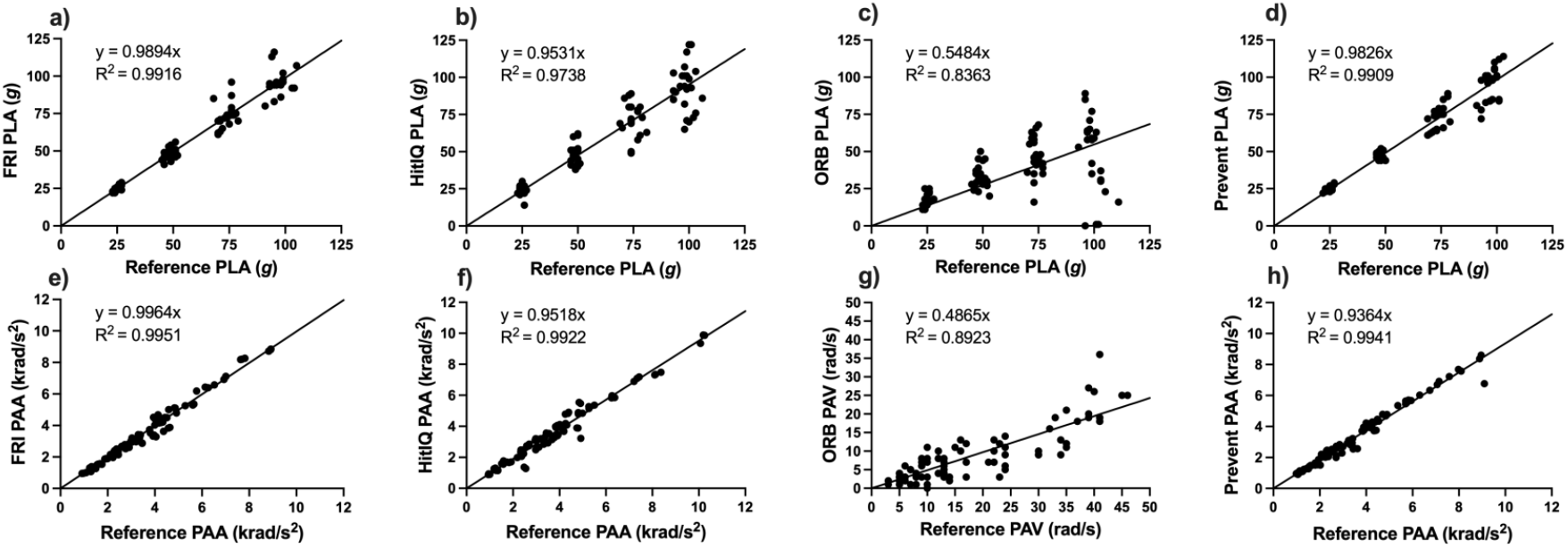
Linear (a) FRI, b) HitIQ, c) ORB, d) Prevent) and angular (e) FRI, f) HitIQ, g) ORB, h) Prevent) kinematic values for each iMG and reference measurements during laboratory bareheaded dummy headform impacts.

**Figure 3.**
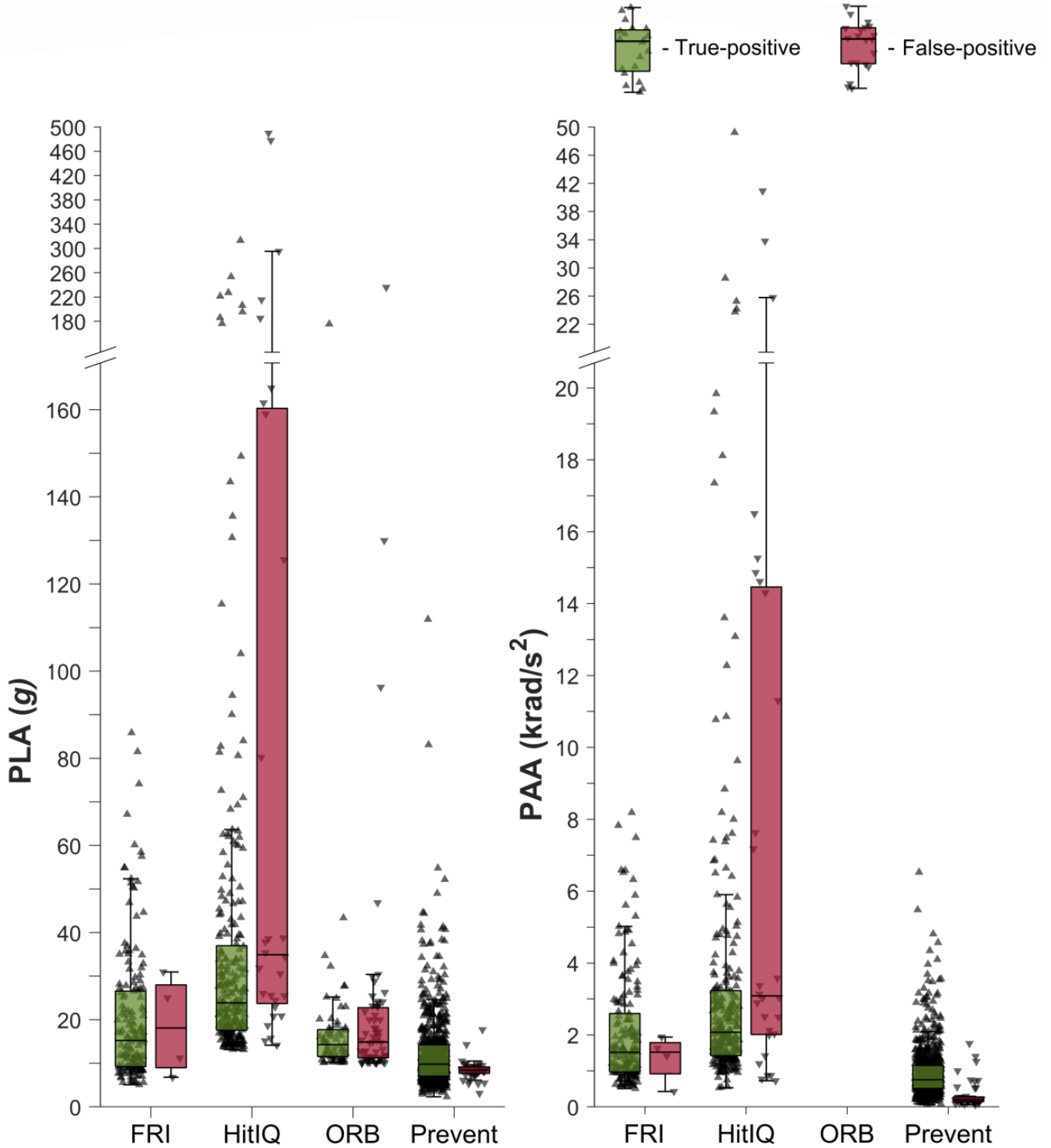
Linear and angular on-field kinematic values from true-positives and false-positives for each iMG from Phase 2. Note ORB did not provide PAA, thus not presented.

## DISCUSSION

This study was the first to determine the validity and feasibility of current iMGs and associated systems, which is vital to consider the error associated with HAE data and maximise the adoption of iMG systems in sport. In Phase 1, FRI, HitIQ, ORB and Prevent had total CCC of 98.3%, 95.3%, 42.5% and 97.9% when compared with dummy headform reference measurements.[2] False-positive (PPV) and false-negative performance (sensitivity) should be considered simultaneously in Phase 2. Overall, Prevent had the highest performance with a PPV of 0.94 and sensitivity of 0.75. For FRI and HitIQ, false-positive performance was higher (PPV = 0.98 and 0.90, respectively) than false-negative performance (sensitivity = 0.51 and 0.40, respectively). ORB had a higher false-negative (sensitivity = 0.75), but lower false-positive (PPV 0.53) performance. To understand likely iMG adoption, the feasibility of current iMG systems were determined. In Phase 3, players reported HitIQ and Prevent to have similar, and FRI and ORB lower ratings for *fit, comfort* and *function*. Practitioners reported the SUS to be highest for Prevent (>80), followed by HitIQ (>70) and FRI (>50). This study demonstrates differences between current iMG systems exist. These findings can be used by sporting organisations to determine the appropriateness of iMG systems, and provides reference data to improve the validity and feasibility of iMG systems, optimising their adoption in sport, supporting player welfare initiatives directly related to long-term brain health.

For Phase 1, FRI, HitIQ, and Prevent had total CCC values >95%, higher than other wearable head sensors previously evaluated.[2] All iMGs achieved higher CCC for PAA than PLA (Table 3). Varying in-laboratory iMG performance has previously been observed (e.g., mean error of 32.4% for PLA[3]) which was attributed to a lack of filtering of iMG kinematic data.[12] This highlights the importance of ongoing hardware/firmware/data-processing development to improve the validity of kinematic data, but also poses a challenge for researchers and practitioners if proprietary algorithms are frequently updated exclusive of independent validation.[13] As such, this study should be repeated, given the rapid development of existing, and emergence of new iMG systems.

Phase 2 evaluated the on-field validity of iMGs (PPV; true-positives and false-positives and sensitivity; true-positives and false-negatives). Prevent had the highest concurrent false-positive and false-negative performance. Both FRI and HitIQ demonstrated high false-positive, but lower false-negative performance, and ORB demonstrated high false-negative, but lower false-positive performance. The different performance by iMGs may be due to the trigger and recording thresholds set by iMG companies, and/or HAE detection algorithms. Prevent had the lowest trigger threshold (8 *g*), *vs*. FRI, ORB (both 10 *g*) and HitIQ (13 *g*), which likely improved false-negative performance when compared to the other iMGs. Lower trigger thresholds could result in more false-positives (due to lower-magnitude iMG-triggered non-HAE being captured) and fewer false-negatives. Most false-positives observed for Prevent (78%) occurred whilst players were running (Supplementary Table 1). These may represent true head accelerations from inertial loading during non-contact events,[14,15] necessitating HAE detection algorithms to differentiate contact from non-contact events.[1] This study defined true-positives as HAE during contact events, however HAE from non-contact events (e.g., running and jumping) may also be desirable in future studies. Further analysis of false-negatives for FRI revealed that the iMG algorithm incorrectly binned 17.4% of false-negatives. The FRI algorithm was trained on American Football data,[16] therefore optimising the algorithm using non-helmeted sports would likely improve false-negative performance. HitIQ false-negative performance was likely due to the 13 *g* recording threshold, which was different to the trigger threshold (Table 1). Further analysis of HitIQ data revealed that 42.5% of false-negatives were collected as iMG-triggered events, but subsequently binned as they fell below the 13 *g* recording threshold, and 1.4% of false-negatives were greater than the recording threshold but were misclassified by the iMG algorithm (Supplementary Table 1). Decreasing the 13 *g* recording threshold (or removing it entirely) may improve HitIQ’s false-negative performance, however the effect this would have on false-positive performance is unknown. Increasing the ability to remove false-positives would improve the overall performance of ORB.

When off-camera and inactive assumed false-positives are considered, false-positive performance reduced for all iMGs (FRI, PPV = 0.98 to 0.81; HitIQ, PPV = 0.90 to 0.60; ORB, PPV = 0.53 to 0.37; Prevent, PPV = 0.94 to 0.89; Supplementary Figure 1). No inactive assumed false-positives were observed for Prevent (Supplementary Table 1) which may be due to proximity sensors embedded within Prevent iMGs, allowing for iMG-triggered events which occurred whilst the iMG was not on the players’ teeth to be removed by the iMG. HitIQ and ORB accumulated more off-camera assumed false-positives, and HitIQ and FRI accumulated more assumed false-positives during inactive periods (93.5% of these for FRI came from a single iMG, which may have been faulty; Supplementary Table 1 and 2). Therefore video-verification or alignment to another data-source (e.g., time motion analysis of match events [17]) may be required prior to use in research and/or practice.

Validating the kinematics of on-field HAE is beyond the scope of this study, due to a lack of an accurate reference sensor. Whilst FRI, HitIQ and Prevent all had similar kinematics CCC in Phase 1 (Table 3 and Figure 2), the kinematics between iMGs from Phase 2 varied and some exceeded the 100 *g* magnitude used in Phase 1. Whilst median kinematic data are influenced by different thresholds, the number and range of high magnitude true-positives differ (Figure 2). The number of true-positives with a PLA above 100 and 200 *g*, were zero and zero for FRI, 14 and five for HitIQ, one and zero for Prevent and one and zero for ORB. Similarly, for true-positives which had a PAA above 10 and 20 krad/s^2^, HitIQ reported 14 and five, whilst FRI, ORB and Prevent all reported none. Measuring the kinematics of on-field HAE is associated with numerous challenges including mandible action (e.g., biting), adherence to teeth and vocalisation, which may affect the magnitude of peak kinematics by introducing noise into iMG signals. It is outside of the scope of this study to evaluate the on-field HAE kinematics, however future studies should investigate the kinematic traces to evaluate signal noise of on-field events.

To optimise iMG adoption, Phase 3 and 4 evaluated the feasibility from a player and practitioner perspective. Most players perceived no issues with the iMGs for *fit* (mean response of ‘no’ = 62 to 88%). Individual questionnaire items revealed players perceived FRI and ORB ‘too bulky’ (84% and 79% of players), with fewer expressing this perception for HitIQ and Prevent (37% and 33% of players; Supplementary Table 3). HitIQ and Prevent (both 8/10) had higher median scores for *comfort*, than FRI and ORB (6/10, 5/10; Table 2). In a previous study,[5] a mouthguard intentionally containing common design faults had a median *comfort* rating of 6/10, which can be used as a comparison. For FRI, HitIQ and Prevent, most players (55 to 67%) perceived no issues with the *function* of iMGs. This was lower for ORB (43%; Supplementary Table 3), where 50% of players perceived it interfered with speech ‘a lot’ in comparison to FRI (26%), HitIQ (11%), and Prevent (17%). As such, the design of iMGs (including the housing and positioning of component parts) is a key consideration for companies when considering the adoption of iMGs within sport.

Phase 4 evaluated the usability for FRI, HitIQ and Prevent system preparation and data management. The industry standard of average SUS for internet-based web pages and applications is 68, which both HitIQ (>70) and Prevent (>80) surpassed. FRI was lower (>50), which could influence adoption by practitioners. Both the system preparation and data management scores were similar within iMGs, which may suggest companies have prioritised these areas within their product development or indicate practitioners do not differentiate between these two aspects. The findings of Phase 3 and 4 provide comparative data for iMG evaluation and development, and potential system adoption.

iMGs have the potential to enhance clinical care via the live (where possible) and retrospective monitoring of HAEs. Both the frequency and magnitude of HAEs that athletes experience within a match, training week, over a season and career can be used to inform player welfare initiatives. Practically, iMGs have the potential to contribute to concussion detection systems, contact load monitoring and return to play practices, which are all priorities in sport.

### Limitations

This study is not without its limitations. Whilst peak resultant kinematic values are important, so are other signal characteristics such as pulse-time and frequency content, not assessed in Phase iMG companies determined their own recording or trigger thresholds, influencing Phase 2 performance. It is unclear if false-negatives associated with no iMG-triggered event were due to one-on-one shoulder tackles not exceeding trigger thresholds or whether iMGs did not capture HAE events for another reason. iMGs should record data from all contact events, regardless of magnitude to gain a robust understanding of player HAE. Only one camera view was available for Phase 2, thus some iMG-triggered events could not be verified. Given the camera followed the ball, and every tackle, these iMG-triggered events were assumed false-positives and not the result of contact events. Future studies should use multiple camera angles to capture all iMG-triggered events during match-play. Not all iMG companies used automatic data processing pipelines for Phase 1 and 2 which may be advantageous for larger-scale project given the amount of data produced (Table 1), and was not assessed in Phase 4. Finally, to allow iMG companies to optimise algorithms and re-test performance, and to consider new iMGs this study should be repeated. To satisfy this point, it is proposed that this protocol is repeated with data collection starting in April 2022.

In conclusion, for the first time this study evaluated the validity and feasibility of current iMG systems, which have potential to provide data on HAE, and inform player welfare initiatives directly related to long-term brain health. This study showed that when compared with reference measurements from a dummy headform, FRI, HitIQ, Prevent had lower, and ORB had higher measurement error. When video-verification was used to determine on-field validity, Prevent had the highest false-negative performance, and second-highest false-positive performance. FRI and HitIQ had higher false-positive performance than false-negative performance, and ORB had higher false-negative performance than false-positive performance. To support the adoption of the iMG systems by players and practitioners, feasibility ratings were provided. HitIQ and Prevent achieved similar high ratings for *fit, comfort* and *function*, and practitioners reported the feasibility of iMG systems to be highest for Prevent, followed by HitIQ, then FRI. For the first time, this study provides data on the strengths and limitations of iMG systems. This can be used by sporting organisations and provides reference data for iMG systems to optimize hardware and software, to improve both the validity of iMG data and feasibility of iMG systems from a player and practitioner perspective, optimizing the adoption of iMGs in sport.

## Data Availability

All data produced in the present work are contained in the manuscript

## Funding

No funding was received to undertake this study.

## Competing interests

GT, JT and DW have received Prevent Biometrics iMGs free of charge for use with Leeds Rhinos RLFC, during training and matches for a research project. BJ, MAD, KT are employed by Leeds Rhinos in a consultancy capacity. GT, JT and EF use Prevent Biometrics iMG for research projects funded by World Rugby. EF is employed by World Rugby. BJ, JT, DW, KT, CO, KS, GP, SH, MAD, GT are involved in the Rugby Football League TaCKLE (Tackle and Contact Kinematic Load & Expsoure) project. BJ and KT will be involved in the procurement of iMGs for the TaCKLE project. BJ, GP are employed in a consultancy capacity by the Rugby Football League. KS is employed by the Rugby Football Union. MB and SR lead the Virginia Tech Helmet Lab. Leeds Rhinos, World Rugby and Virginia Tech are research partners of Prevent Biometrics.

## Acknowledgements

The authors would like to thank all players and staff at Hull KR RLFC, Leeds Rhinos RLFC, St Helens RLFC, Wakefield Trinity RLFC, Warrington Wolves RLFC, and Wigan Warriors RLFC for participating in the study. The authors would like to acknowledge the funding (dental scans, project expenses and Virginia Tech lab costs) provided by Leeds Beckett University, Carnegie School of Sport. The authors would like to thank all Super League CEOs, Laura Fairbank and Karen Moorhouse (Rugby Football League) for supporting the project. The authors acknowledge the support of Brianna Mulhern and Mily Spiegelhalter for supporting data collection. The authors would like to thank Dr Anthony Lovat (OPRO) for reviewing dental scans. Finally, the authors would like to acknowledge their sincere gratitude to Dr Nate Dau and Dr Lee Gabler (Biocore [Football Research Inc; FRI]), Damien Hawes and Tom Laudenbach (HitIQ), Robert Paterson and Thomas Quinn (ORB innovations), and Drew Goodger and Dr Adam Bartsch (Prevent Biometrics) for their support and cooperation during the study.

## Ethical approval

This project was approved by Leeds Beckett University, Local Ethics Committee (85551).

**Supplementary Table 1.**
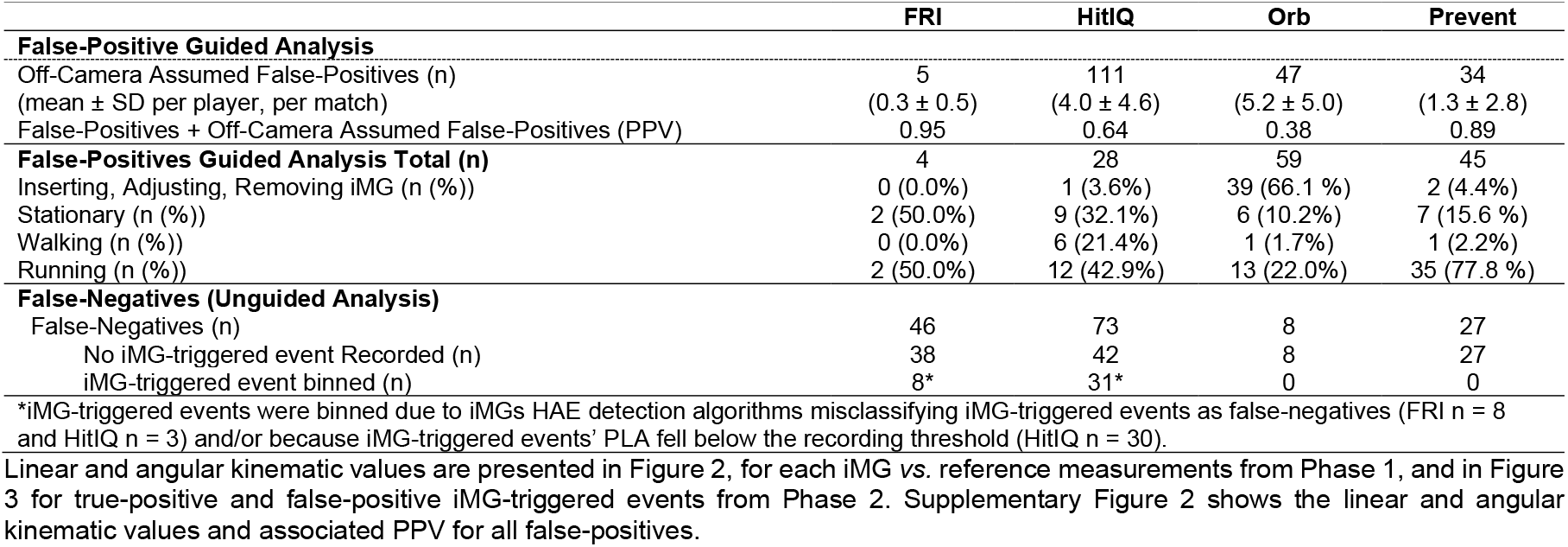
Breakdown of Assumed False-Positives, PPV for Off-Camera Assumed False-Positives, and Breakdown of False-Negatives from Phase 2.

**Supplementary Table 2.**
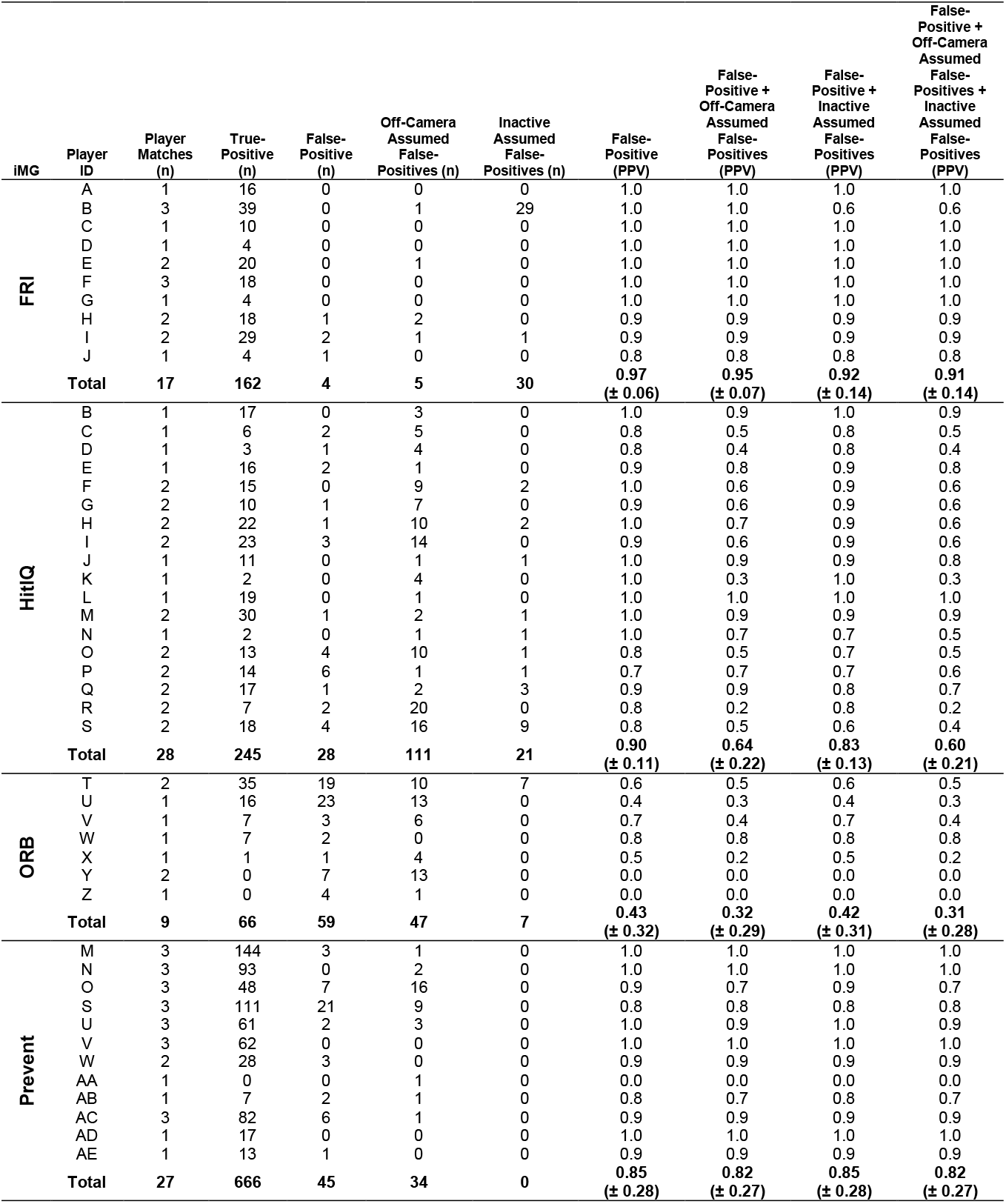
True-Positives, False-Positives and associated PPV for each player from Phase 2.

| iMG | Player ID | Player Matches (n) | True-Positive (n) | False-Positive (n) | Off-Camera Assumed False-Positives (n) | Inactive Assumed False-Positives (n) | False-Positive (PPV) | False-Positive + Off-Camera Assumed False-Positives (PPV) | False-Positive + Inactive Assumed False-Positives (PPV) | False-Positive + Off-Camera Assumed False-Positives + Inactive Assumed False-Positives (PPV) |
| --- | --- | --- | --- | --- | --- | --- | --- | --- | --- | --- |
| FRI | A | 1 | 16 | 0 | 0 | 0 | 1.0 | 1.0 | 1.0 | 1.0 |
|  | B | 3 | 39 | 0 | 1 | 29 | 1.0 | 1.0 | 0.6 | 0.6 |
|  | C | 1 | 10 | 0 | 0 | 0 | 1.0 | 1.0 | 1.0 | 1.0 |
|  | D | 1 | 4 | 0 | 0 | 0 | 1.0 | 1.0 | 1.0 | 1.0 |
|  | E | 2 | 20 | 0 | 1 | 0 | 1.0 | 1.0 | 1.0 | 1.0 |
|  | F | 3 | 18 | 0 | 0 | 0 | 1.0 | 1.0 | 1.0 | 1.0 |
|  | G | 1 | 4 | 0 | 0 | 0 | 1.0 | 1.0 | 1.0 | 1.0 |
|  | H | 2 | 18 | 1 | 2 | 0 | 0.9 | 0.9 | 0.9 | 0.9 |
|  | I | 2 | 29 | 2 | 1 | 1 | 0.9 | 0.9 | 0.9 | 0.9 |
|  | J | 1 | 4 | 1 | 0 | 0 | 0.8 | 0.8 | 0.8 | 0.8 |
| Total |  | 17 | 162 | 4 | 5 | 30 | 0.97<br>(± 0.06) | 0.95<br>(± 0.07) | 0.92<br>(± 0.14) | 0.91<br>(± 0.14) |
| HitIQ | B | 1 | 17 | 0 | 3 | 0 | 1.0 | 0.9 | 1.0 | 0.9 |
|  | C | 1 | 6 | 2 | 5 | 0 | 0.8 | 0.5 | 0.8 | 0.5 |
|  | D | 1 | 3 | 1 | 4 | 0 | 0.8 | 0.4 | 0.8 | 0.4 |
|  | E | 1 | 16 | 2 | 1 | 0 | 0.9 | 0.8 | 0.9 | 0.8 |
|  | F | 2 | 15 | 0 | 9 | 2 | 1.0 | 0.6 | 0.9 | 0.6 |
|  | G | 2 | 10 | 1 | 7 | 0 | 0.9 | 0.6 | 0.9 | 0.6 |
|  | H | 2 | 22 | 1 | 10 | 2 | 1.0 | 0.7 | 0.9 | 0.6 |
|  | I | 2 | 23 | 3 | 14 | 0 | 0.9 | 0.6 | 0.9 | 0.6 |
|  | J | 1 | 11 | 0 | 1 | 1 | 1.0 | 0.9 | 0.9 | 0.8 |
|  | K | 1 | 2 | 0 | 4 | 0 | 1.0 | 0.3 | 1.0 | 0.3 |
|  | L | 1 | 19 | 0 | 1 | 0 | 1.0 | 1.0 | 1.0 | 1.0 |
|  | M | 2 | 30 | 1 | 2 | 1 | 1.0 | 0.9 | 0.9 | 0.9 |
|  | N | 1 | 2 | 0 | 1 | 1 | 1.0 | 0.7 | 0.7 | 0.5 |
|  | O | 2 | 13 | 4 | 10 | 1 | 0.8 | 0.5 | 0.7 | 0.5 |
|  | P | 2 | 14 | 6 | 1 | 1 | 0.7 | 0.7 | 0.7 | 0.6 |
|  | Q | 2 | 17 | 1 | 2 | 3 | 0.9 | 0.9 | 0.8 | 0.7 |
|  | R | 2 | 7 | 2 | 20 | 0 | 0.8 | 0.2 | 0.8 | 0.2 |
|  | S | 2 | 18 | 4 | 16 | 9 | 0.8 | 0.5 | 0.6 | 0.4 |
| Total |  | 28 | 245 | 28 | 111 | 21 | 0.90<br>(± 0.11) | 0.64<br>(± 0.22) | 0.83<br>(± 0.13) | 0.60<br>(± 0.21) |
| ORB | T | 2 | 35 | 19 | 10 | 7 | 0.6 | 0.5 | 0.6 | 0.5 |
|  | U | 1 | 16 | 23 | 13 | 0 | 0.4 | 0.3 | 0.4 | 0.3 |
|  | V | 1 | 7 | 3 | 6 | 0 | 0.7 | 0.4 | 0.7 | 0.4 |
|  | W | 1 | 7 | 2 | 0 | 0 | 0.8 | 0.8 | 0.8 | 0.8 |
|  | X | 1 | 1 | 1 | 4 | 0 | 0.5 | 0.2 | 0.5 | 0.2 |
|  | Y | 2 | 0 | 7 | 13 | 0 | 0.0 | 0.0 | 0.0 | 0.0 |
|  | Z | 1 | 0 | 4 | 1 | 0 | 0.0 | 0.0 | 0.0 | 0.0 |
| Total |  | 9 | 66 | 59 | 47 | 7 | 0.43<br>(± 0.32) | 0.32<br>(± 0.29) | 0.42<br>(± 0.31) | 0.31<br>(± 0.28) |
| Prevent | M | 3 | 144 | 3 | 1 | 0 | 1.0 | 1.0 | 1.0 | 1.0 |
|  | N | 3 | 93 | 0 | 2 | 0 | 1.0 | 1.0 | 1.0 | 1.0 |
|  | O | 3 | 48 | 7 | 16 | 0 | 0.9 | 0.7 | 0.9 | 0.7 |
|  | S | 3 | 111 | 21 | 9 | 0 | 0.8 | 0.8 | 0.8 | 0.8 |
|  | U | 3 | 61 | 2 | 3 | 0 | 1.0 | 0.9 | 1.0 | 0.9 |
|  | V | 3 | 62 | 0 | 0 | 0 | 1.0 | 1.0 | 1.0 | 1.0 |
|  | W | 2 | 28 | 3 | 0 | 0 | 0.9 | 0.9 | 0.9 | 0.9 |
|  | AA | 1 | 0 | 0 | 1 | 0 | 0.0 | 0.0 | 0.0 | 0.0 |
|  | AB | 1 | 7 | 2 | 1 | 0 | 0.8 | 0.7 | 0.8 | 0.7 |
|  | AC | 3 | 82 | 6 | 1 | 0 | 0.9 | 0.9 | 0.9 | 0.9 |
|  | AD | 1 | 17 | 0 | 0 | 0 | 1.0 | 1.0 | 1.0 | 1.0 |
|  | AE | 1 | 13 | 1 | 0 | 0 | 0.9 | 0.9 | 0.9 | 0.9 |
| Total |  | 27 | 666 | 45 | 34 | 0 | 0.85<br>(± 0.28) | 0.82<br>(± 0.27) | 0.85<br>(± 0.28) | 0.82<br>(± 0.27) |

**Supplementary Table 3.** *Fit, Comfort* and *Function* for each questionnaire item from Phase 3.

| Question | FRI | HitIQ | Orb | Prevent |
| --- | --- | --- | --- | --- |
| ***Fit*** |  |  |  |  |
| ***(% of participants who responded 'no')*** |  |  |  |  |
| <sup>a</sup>Too loose | 95 | 89 | 78 | 94 |
| <sup>a</sup>Too tight | 84 | 95 | 67 | 83 |
| <sup>a</sup>Too bulky | 16 | 63 | 22 | 67 |
| <sup>a</sup>Too small or thin | 100 | 95 | 94 | 100 |
| <sup>a</sup>Held your mouth open | 42 | 79 | 39 | 72 |
| <sup>a</sup>Held your teeth apart | 89 | 89 | 72 | 89 |
| <sup>a</sup>Created pain in your jaw muscles | 95 | 95 | 83 | 94 |
| <sup>a</sup>Created an uneven bite | 95 | 100 | 67 | 83 |
| ***Comfort*** |  |  |  |  |
| ***(Median [IQR] of responses to a 0-10 likert scale)*** |  |  |  |  |
| <sup>b</sup>Overall comfort | 6 (5,7) | 7 (6,8) | 3 (2,5) | 7 (5,8) |
| <sup>b</sup>Lip comfort | 5 (4,6) | 7 (6,8) | 5 (3,6) | 8 (7,8) |
| <sup>b</sup>Gum comfort | 6 (5,7) | 7 (6,8) | 5 (2,6) | 7 (7,8) |
| <sup>b</sup>Tongue comfort | 7 (6,8) | 8 (7,8) | 5 (3,5) | 8 (7,8) |
| <sup>b</sup>Tooth comfort | 6 (6,8) | 8 (7,9) | 5 (3,7) | 8 (5,8) |
| <sup>b</sup>How well it held in your mouth | 6 (6,8) | 8 (8,9) | 5 (3,7) | 8 (7,9) |
| <sup>b</sup>Ease of breathing | 6 (5,7) | 7 (6,8) | 5 (3,6) | 8 (6, 8) |
| ***Function*** |  |  |  |  |
| ***(% of participants who responded 'no', 'a little', 'a lot')*** |  |  |  |  |
| <sup>c</sup>Interferes with speech | 16, 58, 26 | 21, 68, 11 | 11, 39, 50 | 44, 39, 17 |
| <sup>c</sup>Interferes with swallowing | 58, 42, 0 | 58, 42, 0 | 28, 44, 28 | 56, 44, 0 |
| <sup>c</sup>Causes a dry mouth | 47, 42, 11 | 69, 26, 5 | 50, 33, 17 | 72, 22, 6 |
| <sup>c</sup>Causes nausea | 100, 0, 0 | 95, 5, 0 | 83, 6, 11 | 94, 6, 0 |
| Mean 'No' | 55.3 | 60.8 | 43 | 66.5 |
| Mean 'A little' | 35.3 | 35.3 | 30.5 | 27.8 |
| Mean 'A lot' | 9.3 | 4.0 | 26.5 | 5.8 |

<sup>a</sup> With regards to the fit of the mouthguard, did you feel that this mouthguard was; <sup>b</sup> On reflection following the training sessions, how did you rate the comfort of this mouthguard for the following elements?; <sup>c</sup> On reflection following the training sessions, how did you rate the function of this instrumented mouthguard for the following elements?

**Supplementary Figure 1.**
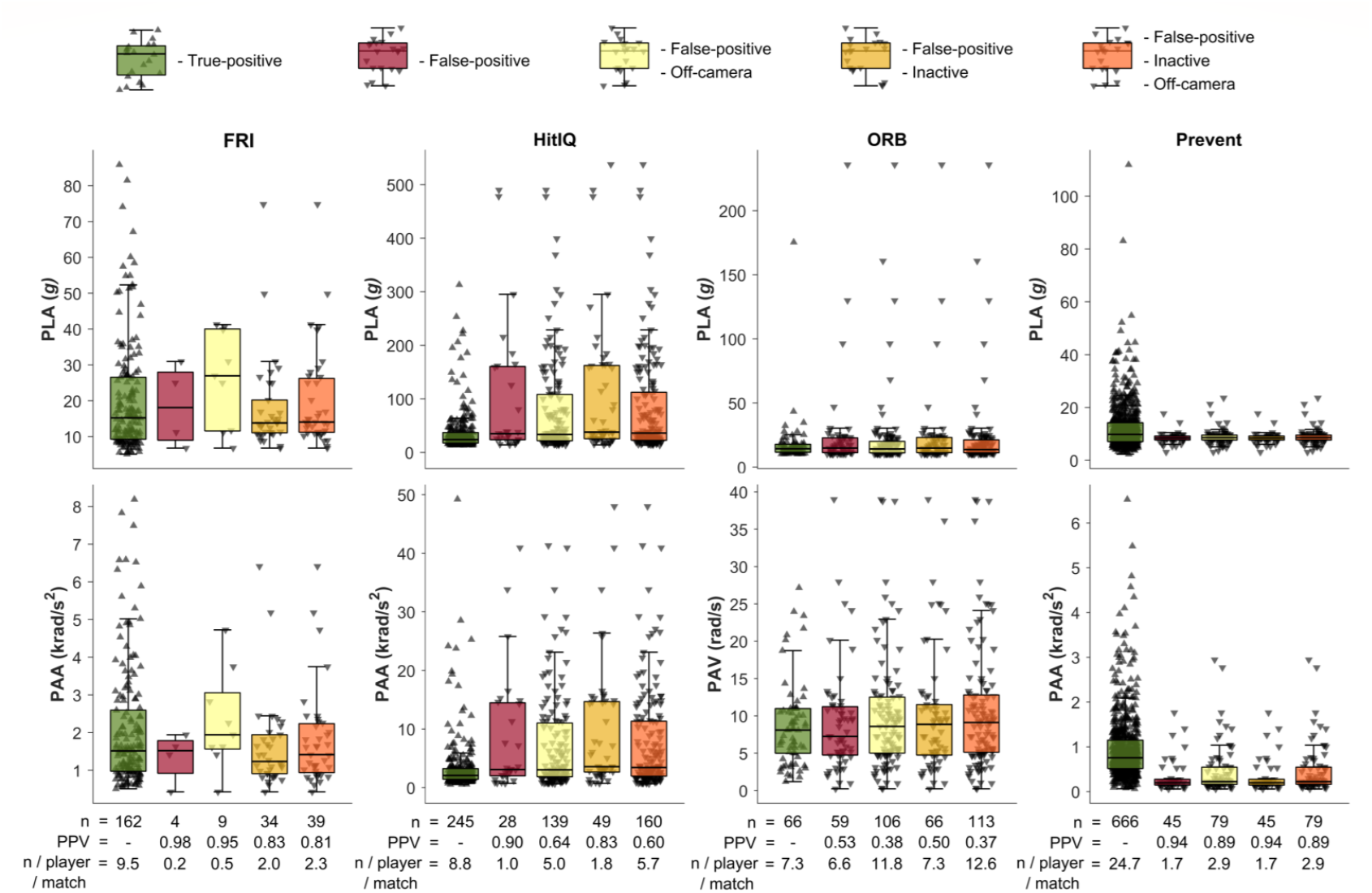
Peak linear and angular on-field kinematic values, associated PPVs, number per player, per match of true-positives, false-positives, and combinations of false-positives and assumed false-positives for each iMG from Phase Note. Y axis scale specific to data reported by each iMG.

## Notes

### Funding Statement

This study did not receive any funding

### Author Declarations

Ethical approval: This project was approved by Leeds Beckett University, Local Ethics Committee (85551).

